# PREVALENCE AND HISTOLOGICAL PATTERNS OF PROSTATE CANCER AMONG PATIENTS PRESENTING WITH OBSTRUCTIVE LOWER URINARY TRACT SYMPTOMS AT MBARARA REGIONAL REFERRAL HOSPITAL

**DOI:** 10.1101/2023.07.25.23293158

**Authors:** Willy Kyegombe, Epodoi Joseph, Okidi Ronald, Kitara Lagoro, Ekuk Eddymond, Nimusima Aniitah, Marvin Mutakooha Mwesigwa

**Author notes:** **Author Contributions** **Willy Kyegombe**: Principal investigator, Patient management and care; Manuscript drafting and review. **Dr Marvin Mutakooha Mwesigwa MD, FCS Urology**: Supervision of protocol writing and execution, Patient management and care; Manuscript drafting and Review. **Dr Epodoi Joseph MD FCS Urology**: Supervision of protocol writing and execution, Patient management and care; Manuscript drafting and Review. **Dr Okidi Ronald MD,FCS Urology**: Protocol writing and execution, Manuscript drafting and Review. **Prof Kitara Lagoro**: Protocol writing and execution, Manuscript drafting and Review. **Ekuk Eddymond**: Protocol writing and execution, Manuscript drafting and Review **Nimusima Anitah**: Protocol writing, data processing, Manuscript drafting and Review. **Funding Statement** This study was executed with funding from the First mile scholarship program.

## Abstract

**Background:** Prostate cancer is currently the second commonest male cancer in Uganda. Despite this, men are more likely to be tested for prostate cancer only after presenting with obstructive Lower urinary tract symptoms [1] since these are a common presenting complaint among elderly males [2]. However, there is still a lack of published local information on the clinical and biochemical patterns of prostate cancer among patients with lower urinary tract symptoms in Uganda which can negatively affects the equitable distribution of resources for cancer care.

**Aims:** To determine the Patterns of PSA, DRE, histological findings, and prevalence of prostate cancer among patients presenting to Mbarara regional referral hospital with obstructive LUTS.

**Materials and Methods:** A cross-sectional study of 140 patients with obstructive LUTS. Demographics, presenting symptoms were collected using a structured questionnaire and IPSS index, followed by total serum prostate specific antigen level estimation and Digital rectal examination. Patients who had a total serum Prostate Specific Antigen level above 4ng or an abnormal DRE underwent digitally guided transrectal prostate biopsy.

**Results:** Majority had severe LUTS (n=103,73.57%) with a median tPSA of 14.4 ng/ml and met the criteria for the digitally guided transrectal trucut prostate biopsy(n=99,70.71%). DRE abnormalities were found in 57.14% n=80 of the participants. The commonest abnormalities were prostatic enlargement (n=76,54.29%), hard prostatic consistency (29.2% n=41), loss of the median groove (35.71% n= 50) and nodular prostate (n=60, 42.86%). The prevalence of prostate cancer was high at 52.21% (59/11395% C.I:30.1-46.3), and over 93.3% of the cancer postive patients exhibited abnormalities on DRE.

Prostate adenocarcinoma was the only histological type in all biopsies. The mean Gleason score was 8 (±1.148) and the majority had a Gleason score of 8 and above. (77.78%, n=35).

**Conclusions:** The prevalence of prostate cancer among men presenting to Mbarara Regional referral hospital with obstructive LUTS was high and majority of these had poorly differentiated prostate adenocarcinoma.

## Introduction

World over, prostate cancer is currently ranked as the second commonest male cancer[3] with over seventy five percent of the cases being observed among men over the age of sixty five(Parkin et al., 2001). African statistics are currently also indicating an upward trend in prostate cancer prevalence and incidence, with the main drivers of this being, aging population growth together with the associated increase in risk factors, [4, 5].

In Uganda prostate cancer is the second commonest type of male cancer with a current age standardized incidence rate of 39.6 per 100,000[6] and one year prevalence of 28.1%, figures which are currently higher than what is reported by most African countries. Unfortunately, over 75% of cases are always advanced and incurable at first presentation[1, 7]. Usually early prostatic cancer is asymptomatic, as it progresses patients can develop, heamatura and heamatochezia and then in distant metastatic disease can a have a variety of symptom ranging from back pain, pathological fractures, and respiratory symptoms to severe aneamia [1]. Among all these symptoms LUTS are generally becoming more prevalent among patients with confirmed prostate cancer [1, 8-10]. This further asserts findings by Hamilton and sharp that locally advanced cancer can cause LUTS that are similar to those for benign prostatic hypertrophy [11] .In addition to the LUTS locally advanced prostate cancer can present with unique DRE findings like fixation of rectal skin, bleeding per rectum, increased nodularity. [12]. A large number of males are currently presenting with lower urinary tract symptoms[2] and are more likely to have their fast test for prostate cancer at this time [7] despite of the current lack of global consensus on the relation between LUTS and prostate cancer. This makes it imperative to ascertain the local and regional clinical and biochemical profile of prostate cancer in this population of patients. Currently however information still not widely available in our setting yet it will be crucial in successfully developing strategic plans to combat the current late presentation with advanced prostate cancer and guide equitable distribution of cancer care resources. Therefore this study, described the commonest DRE findings and PSA patterns, prostate cancer prevalence together with a description of the commonest histological types and gleason scores in a small population of men presenting with LUTS to regional referral hospital in western Uganda

## Materials and methods

This a cross-sectional descriptive study of 140 men aged 50 years and above who presented to Mbarara Regional Referral Hospital with lower Urinary Tract symptoms. Data on the severity of the LUTs was collected using the AUS International prostate symptom score, a blood sample for total serum prostate specific antigen estimation was obtained from each patient before undergoing a digital rectal examination. Patients who had an abnormality in either or both Digital rectal examination or total serum Prostate specific antigen underwent a trucut prostate biopsy for histological analysis from Mbarara University Department of Pathology. Data analysis was done with STATA 13.0 and using descriptive statistics results were presented in the form of means for continuous variables, counts were done for categorical variables. Abnormal total serum PSA levels were set at 4ng/ml and above. Abnormal Digital Rectal Examination referred to the Detection of any of the following findings in the prostate gland; enlargement, nodules, induration, asymmetry, loss of the median groove, and tethering of rectal mucosa to the prostate gland.

## Results

Majority of the study participant were aged between 61 to 70 years 32.86% (46/140, mean= 69.8+/-11.1). They came from mainly the districts of Mbarara (30%) and Insingiro (25%). Their major occupation was peasant farming. (n=111, 79.28%). Majority 73.57% (103/140) of them presented with severe LUTS and qualified for a trucut prostate biopsy 70.71(99/140) as shown in figure 1.

**Table 1:**
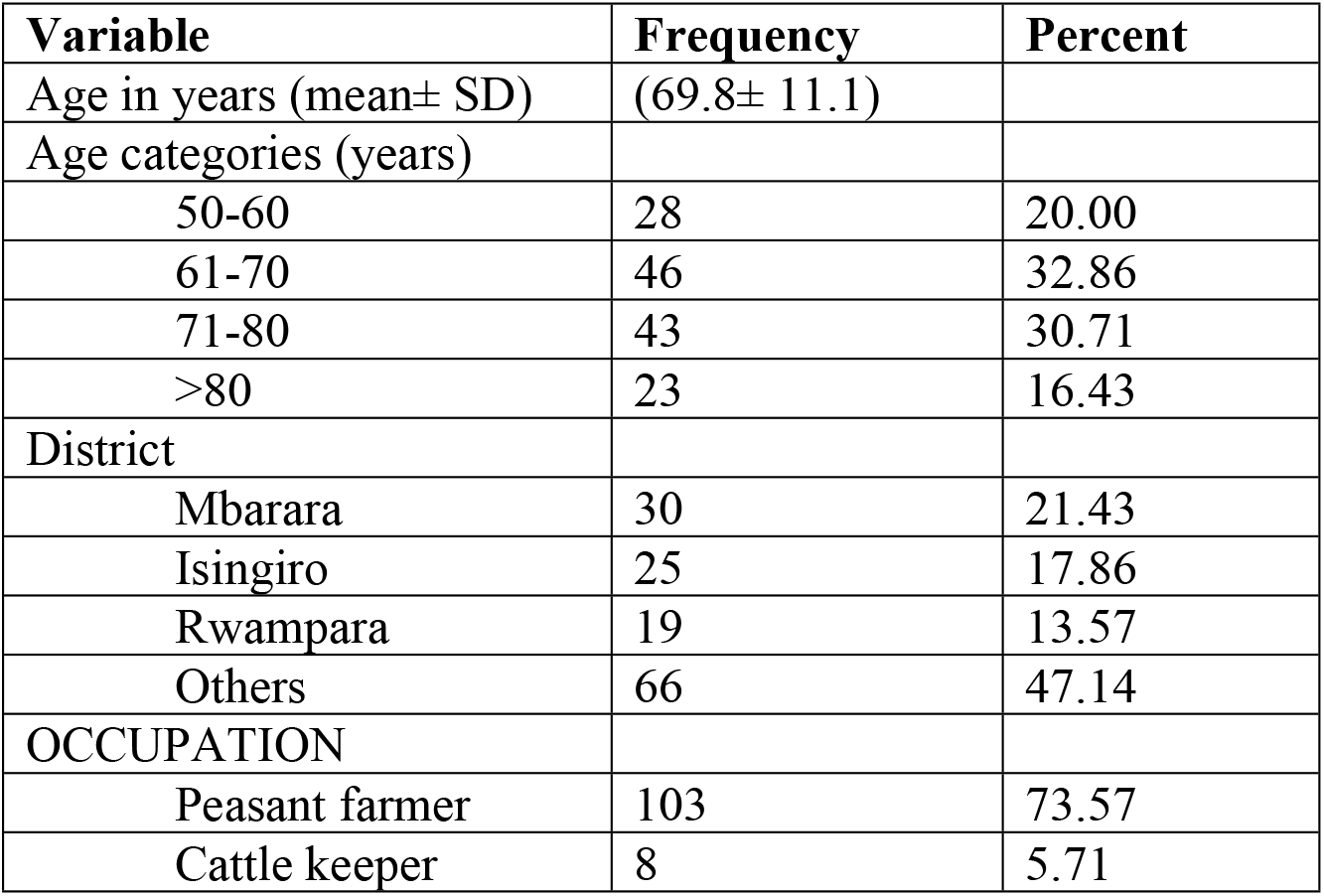
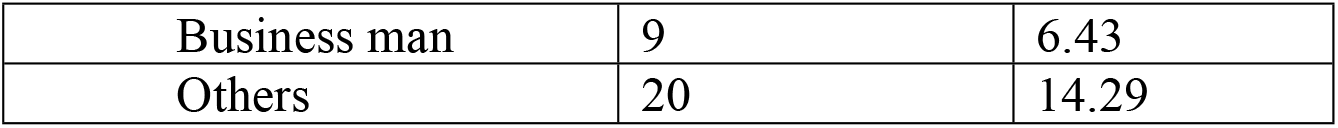
Sociodemographic characteristics of the study participants(n=140)

| Variable | Frequency | Percent |
| --- | --- | --- |
| Age in years (mean± SD) | (69.8± 11.1) |  |
| Age categories (years) |  |  |
| 50-60 | 28 | 20.00 |
| 61-70 | 46 | 32.86 |
| 71-80 | 43 | 30.71 |
| >80 | 23 | 16.43 |
| District |  |  |
| Mbarara | 30 | 21.43 |
| Isingiro | 25 | 17.86 |
| Rwampara | 19 | 13.57 |
| Others | 66 | 47.14 |
| OCCUPATION |  |  |
| Peasant farmer | 103 | 73.57 |
| Cattle keeper | 8 | 5.71 |
| Business man | 9 | 6.43 |
| Others | 20 | 14.29 |

**Figure 1:**
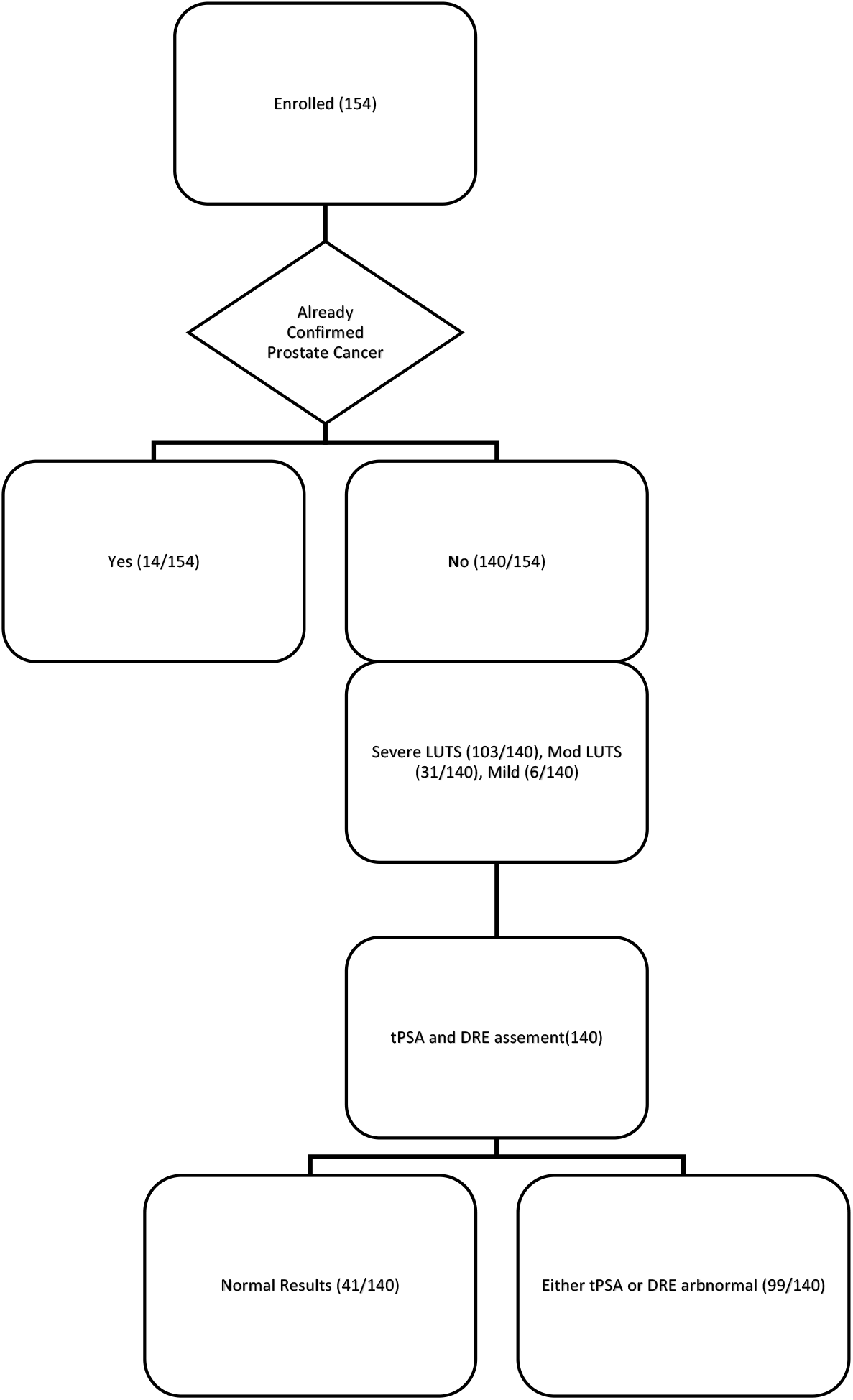
Participant flow chart.

The prevalence of prostate cancer was found to be high at 52.21% (59/113, 95% C.I:30.1-46.3)

The median PSA was 14.3 ng/ml; the lowest PSA value recorded was 0.29ng/ml while the highest was 592ng/ml. Majority of the participants (99/140, 71%) had TPSA values above 4ng/ml. Abnormalities on DRE were found in 57.14%, (80/140) of the participants who presented with LUTS.

However, among patients who had histologically confirmed prostate cancer DRE abnormalities were present in over 93.3% (42/45), with the commonest abnormalities being Nodularity 82.2% (37/45), loss of the median groove 82.2% (37/45), hard prostatic consistency 77.8% (35/45), prostatic enlargement 86.67% (39/45).

**Table 2:** Showing percentage of patients per tPSA catergory.

|  |  |  |
| --- | --- | --- |
| **Median** | 14.33 |  |
| **tPSA categories ng/ml** | **Frequency** |  |
| 0-4 | 41 | **Percent** |
| 4.1-10 | 22 | 29.29% |
| >10 | 77 | 15.71% |
|  |  | 55.00% |

All the patients with histologically confirmed malignancy had prostate adenocarcinoma. The mean Gleason risk score was 8 (±1.148). When the individual Gleason scores were classified into three categories. Majority of the adenocarcinoma had a Gleason score of 8 and above. (77.78%, n=35).

**Table 2:** Showing the distribution of the different Gleason scores.

|  |  |
| --- | --- |
| **Mean** | 8 ($\pm 1.148$) |
| **Gleason score Category** | **Count (Percent)** |
| 2-6 | 5(11.11) |
| 7 | 5(11.11) |
| 8 and above | 35(77.78) |
| **Total** |  |

## Discussion

Majority of our participants were aged between 61 to 70years 32.86% (46/140) with a mean 69.8+/-11.1) this age group encompasses the currently the globally accepted mean age of 68 for diagnosis with prostate cancer.[13] and similar to that reported by other Ugandan studies [1]. This age also coincides the commonest age at presentation with LUTS[2], and the high rates of prostate cancer detected during this study could indicate that clinicians should encourage this population to always screen for prostate cancer once they present with LUTS.

The prevalence of prostate cancer was 52.21% (59/113,), 95% C.I:30.1-46.3). This was higher than that reported by most studies that did not consider the presence of LUTS as a major determinant in the denominator. A Brazilian meta-analysis of Twelve studies comparing the prevalence of prostate cancer among Africans and Caucasian men, revealed an overall prevalence of prostate cancer at 9.6% in Black and 5.6% in White men[14]. A Caribbean screening study of men aged 40 to 79 years that did not include LUTS as part of the screening criterion also found a lower prevalence at 10.7% [15]. However in addition to these studies not including LUTS as prerequisite for inclusion, they were conducted in the community on health men and therefore lacked the backsons bias that can increase prevalence.

In agreement to our findings some meta analytical studies have elucidated that locally advanced cancer could have obstructive Lower Urinary Tract Symptoms that are similar to those for benign prostatic hypertrophy[11] despite several population studies finding no statistically significant relation ship between LUTS and prostate cancer[12, 16].

The median PSA was 14.3 ng/ml, and majority of the participants 71% (99/140), had TPSA values above 4ng/ml. This means that 71 percent of all male patients 50 years and above presenting to OPD with LUTS qualify for further asassessment for prostate cancer.We did not find any local or continental studies that had specifically profiled PSA among patients with LUTS however, retrospective sub-Saharan studies conducted on already confirmed prostate cancer patients reveal higher PSA values and LUTS as the commonest presenting complaints ie median PSA values of 91.3ng/ml [1] in kampala, 92.6ng/ml in Nigeria[10] 96ng/ml in Nigeria[17], whereas the lowest rates of abnormal PSA values are being reported from community based studies like 2.6ng/ml in Brazil and less than 29% with abnormal PSA values in screening study conducted in the Caribbeans [18] This information would be vital in planning the amount of resources required for activities like community out reaches in testing for prostate cancer

Our findings suggest that abnormalities on DRE are common 57.14%, (80/140) among patients with LUTS, and therefore physicians should endeavour to always perform a DRE in this population of patients since also our study found that among patients who had histologically confirmed prostate cancer, over 93.3% (42/45) had exhibited abnormalities on DRE before trucut biopsy was done.

This concurs with another cross sectional study that assessed the clinical relevancy or Digital Rectal Examination in general medical and urology clinics, abnormalities in Digital Rectal Examination were found over 67% of the participants [19], since these patients were recruited during clinics, it could mean that similar to our study population they were already symptomatic at presentation, further highlighting the relevancy of performing a DRE in patients with LUTS. Chiang et al 2015 found that approximately a third of men 50 years and older will have some DRE findings that are due to a prostate pathology [20]. The relevancy of a well performed digital rectal examination had however already been supported by results from autopsy studies that has revealed that 70% of cases of prostate cancer shows local extension which is likely to be detected during DRE [21].

The only histological subtype of prostate cancer found among our study population was adenocarcinoma, other rare histological patterns that account for 5–10% of the carcinomas that originate from the prostate, like small cell, ductal adenocarcinoma, sarcomatoid, basal cell, squamous cell, adenosquamous (ASC), and urothelial carcinoma [22] [23] were not found among any of our participants. [1, 7, 17, 24]

Another retrospective re-examination of prostate specimens collected between 1994 and 2004 at the university teaching hospital of Calabar Nigeria also revealed 99% cases being adenocarcinoma.) with only one mesenchymal tumor (rhabdomyosarcoma) (1%) [25]. This could mean that prevalence of other rare histological subtypes of prostate cancer is even much more unlikely to occur among men of black descent however further evaluation with different study designs and higher sample sizes are still needed to verify this.

The mean Gleason score was high at 8 and when the individual scores were clustered into three risk categories majority of the patients who had prostate cancer (35/45, 77.78%) had Gleason scores above 8 which means that majority of our prostate cancer patients had high risk poorly differentiated cancer that was most likely incurable or presented late with advanced cancers.

This was higher than that detected by Oluwole et al in a 10-year retrospective review of 151 cases at Ahmadu Bello University teaching hospital Zaria Nigeria, 51.6% of the cancers were poorly differentiated and 58.4% moderately differentiated or rather moderate risk. [24]. However, this having been a retrospective review, presentation with LUTS was not a major inclusion criterion.

A study done by Yahaya at Mulago Mulago National Refferal Hospital Uganda in 2019, also revealed that a significant proportion of his patients 44.6% had a Gleason score of ≥ 8 [8] however still lower than the percentage determined by our study

A South African study published in 2014 revealed that black South Africans presented with significantly more aggressive disease defined by Gleason score >7 and poorly differentiated tumor grade within rural versus urban localities [26]. Okuku during his retrospective review at Makerere found that majority of the specimens had high-risk disease with a Gleason score of 9 or 10 in 66.7% of the patients. [1]

A study with predominantly white participants that was conducted in USA Louisiana to assess the pattern of prostate cancer among patients with abnormal Digital Rectal Examination findings and total serum prostate specific antigen levels below 4ng/ml, diagnosed low-risk cancer in 72.8% of the biopsies. [27] which was the opposite of our findings

The above studies([1, 7, 8, 26] confirm with finding from other studies suggesting that Africans are likely to present with aggressive poorly differentiated prostate cancer unlike people from the white races[28], this could be is due to delayed hospital presentation however, African due to high testosterone levels and genetic predisposition are more likely to develop highly aggressive rapidly progressing prostate cancer[28].

## CONCLUSIONS

This study highlights the fact that majority (70.71%) of men above 50 years who present to OPD with LUTS will need a trucut prostate biopsy to rule out prostate cancer and are also likely to have a high prevalence of prostate cancer at 52.21. A large percentage of patients with confirmed prostate cancer 93.3% (42/45) had abnormal DRE findings and also had poor differentiated adenocarcinoma. It is also imperative that Strategies to minimize late presentation with disease are devised and instituted by the health authorities of the districts where our participants came from. Further studies into the staging of disease at presentation and treatment will be useful in guiding the equitable distribution of resources for treatment of this condition.

## Limitations

Use of digitally guided transrectal prostate biopsy. as we could have missed non palpable nodules.The fact that the study was conducted in a hospital setting that always to symptomatic already sick patients could have raised the prevalence.

## Data Availability

The data underlying the results presented in the study are available from

## Acknowledgements

Special thanks to first mile scholarship program for all the rendered to me during my residency.

